# A computational optimization approach for the automatic generation of Gamma Knife radiosurgery treatment plans

**DOI:** 10.1101/2021.08.26.21262676

**Authors:** Matthew R. Walker, Mehrdad Malekmohammadi, Catherine Coolens, Normand Laperriere, Robert Heaton, Ali Sheikholeslami, Mojgan Hodaie

**Author notes:** Authors contributed equally to this work (Matthew R. Walker, Mehrdad Malekmohammadi).

## Abstract

Gamma Knife (GK) radiosurgery is a non-invasive treatment modality which allows single fraction delivery of focused radiation to one or more brain targets. Treatment planning mostly involves manual placement and shaping of shots to conform the prescribed dose to a surgical target. This process can be time consuming and labour intensive. An automated method is needed to determine the optimum combination of treatment parameters to decrease planning time and chance for operator-related error. Recent advancements in hardware platforms which employ parallel computational methods with stochastic optimization schemes are well suited to solving such combinatorial optimization problems efficiently. We present a method of generating optimized GK radiosurgery treatment plans using these techniques, which we name ROCKET (Radiosurgical Optimization Configuration Kit for Enhanced Treatments). Our approach consists of two phases in which shot isocenter positions are generated based on target geometry, followed by optimization of sector collimator parameters. Using this method, complex treatment plans can be generated, on average, in less than one minute, a substantial decrease relative to manual planning. Our results also demonstrate improved selectivity and treatment safety through decreased exposure to nearby organs-at-risk (OARs), compared to manual reference plans with matched coverage. Stochastic optimization is therefore shown to be a robust and efficient clinical tool for the automatic generation of GK radiosurgery treatment plans.

## Introduction

Gamma Knife® (GK; Elekta, Stockholm, Sweden) radiosurgery is a treatment modality within stereotactic neurosurgery which permits single fraction, focused delivery of radiation to the brain. This technique involves the use of gamma radiation using 192 ^60^Co sources which can be precisely focused to irradiate brain targets while sparing nearby healthy tissue. GK is frequently used in the treatment for lesions with high risk of surgical resection, including brain tumors and arteriovenous malformations (1, 2), as well as trigeminal neuralgia(3, 4) and movement disorders(5–7). The key advantage, in addition to its non-invasive profile, is effective control and conformal delivery of radiation while preserving brain function and with fewer post-operative complications compared to microsurgical resection(8–13).

The current clinical paradigm is a forward planning approach which consists of manually placing and shaping shots throughout the target volume to generate a complex dose distribution. This procedure can be time consuming, taking up to several hours per patient, substantially hindering the clinic schedule and straining healthcare resources. Further, operator variability presents the opportunity for subjective errors to occur. Automated inverse planning software (Wizard™ and Lightning™; Elekta, Stockholm, Sweden) is available but, compared to manual planning, may be insufficiently precise for a satisfactory treatment quality without further manual modification(14). Thus, an efficient and robust planning method is needed to automatically generate treatment plans in a short time which satisfy the clinical requirements of sufficient dose to the target while protecting nearby healthy tissue.

GK treatment planning may be framed as a combinatorial optimization problem as a large array of parameters must be considered in order to craft an effective treatment plan. Recent advances in specialized hardware with parallel computational abilities and stochastic optimization algorithms have made it possible to find global solutions to these problems efficiently(15). Currently available platforms are capable of solving these problems at speeds and scales not possible using traditional computational techniques such as simulated annealing or parallel tempering(15, 16). Similar methods have been successfully implemented for treatment optimization in intensity-modulated radiation therapy, which is notable due to its similarities with GK treatment planning(17, 18).

In this paper we present the clinical application of our GK treatment planning method using a stochastic optimization system which we term ROCKET (Radiosurgical Optimization Configuration Kit for Enhanced Treatments). We apply this technique to a retrospective cohort of brain tumor patients and perform a comparison with manually generated treatment plans. Our hypothesis is that our proposed planning method will result in significantly reduced plan generation time with no significant reductions in metrics of treatment plan quality. For the purposes of this study, we have selected a common type of lesion treated with GK. Acoustic neuromas are often treated with GK due to their location near the brainstem and the risks associated with surgical resection. The proximity of key organs-at-risk (OARs) necessitates a high level of precision in treatment planning, thus making acoustic neuroma an ideal model for optimization studies.

## Methods

### Research Subjects

A total of 49 patients (27 female) with acoustic neuroma were randomly sampled through retrospective chart review with approval from the University Health Network Research Ethics Board. Acoustic neuroma is among the most common tumor types treated with GK and is an ideal model for optimization studies due to the proximity of nearby OARs and the wide range of tumor shapes and sizes. Tumor sizes ranged from 0.23 to 11.17cm^3^. All subjects were previously treated with a Leksell Gamma Knife® Perfexion™ (Elekta, Stockholm, Sweden) unit at the Toronto Western Hospital in Toronto, Ontario, Canada using manually generated treatment plans. Given that this is a retrospective study, the time required to generate these archival plans was estimated to require an average of two hours or more, depending on the complexity of the case. All patients underwent 3T magnetic resonance (MR) imaging with contrast enhancement and computed tomography (CT) imaging for targeting guidance and treatment planning. T1-weighted 3D fast spoiled gradient echo (FSPGR) and T2-weighted 3D fast imaging employing steady-state acquisition (FIESTA) images were acquired with 1mm axial slice thickness and in-plane voxel resolution of 0.39mm × 0.39mm. CT images were acquired for each patient after being fitted with a stereotactic frame for targeting accuracy and stability with voxel size = 0.57 mm × 0.57 mm × 1 mm.

Delineation of tumor targets and nearby OARs was performed manually by the treating physicians (neurosurgeon and radiation oncologist) using pre-treatment MR and CT images. These same structure segmentations were used for dose distribution measurements in both optimized and manual reference plans as well as for isocenter position generation prior to optimization of dose distribution. OARs were present in all subjects as determined by the proximity of critical structures to the tumor. These include, but are not limited to, the brainstem, cochlea, optic chiasm, and trigeminal nerve.

Our ROCKET plan generation procedure is divided into two distinct phases: i) isocenter position determination followed by ii) sector size optimization (SSO). In the isocenter positioning phase, we generate the number of shots and coordinates of each shot in order to coarsely cover the target volume using a novel sphere packing (SP) approach. These isocenter positions remain fixed throughout the subsequent optimization phase. During the SSO phase the sector collimation variables and weights are finely tuned to shape each shot.

To investigate the effects of each phase, we compare three different treatment plans: manual forward planning, Fwd-ROCKET, and SP-ROCKET. Manual forward plans are the retrospectively acquired reference plans which were delivered to patients. Fwd-ROCKET plans use the same manually determined isocenters but with our SSO method applied in the second phase. SP-ROCKET plans are entirely automatically generated with SP in the first phase and SSO in the second phase.

### Treatment Plan Evaluation

Clinical treatment plans are generally assessed on their ability to deliver the prescribed dose to the target while minimizing the dose deposited to adjacent healthy tissue. Quantitative metrics commonly used in the GK clinic are coverage, selectivity, gradient index, and beam-on time (BOT)(19). Coverage refers to the portion of the target volume which receives at least the prescription dose or greater. Selectivity refers to the volume of the prescription isodose which coincides with the target volume. Ideally, coverage and selectivity would be as close as possible to 100%. Gradient index refers to the rate at which dose spatially drops off outside of the prescription isodose volume (calculated as the ratio of half the prescription isodose volume to the volume of the full prescription isodose). Gradient index is kept as low as possible, indicating a sharp dose drop-off with little dose leakage into surrounding healthy tissue. BOT, the exposure time required to deliver a treatment, is also a consideration as total treatment time must be tolerable for patients. Explicit optimization of BOT may also enhance treatment efficiency and safety.

While the quantitative metrics discussed above provide the ability to objectively compare different treatment plans, these measures can be difficult to interpret and may not reflect every clinical concern relevant to the case. It is not possible to achieve optimality in each clinical objective in every case as one may negatively impact another; for example, maximizing a target’s prescription dose coverage may exceed the safe dose threshold of a nearby OAR. Further, there is some heterogeneity amongst facilities regarding the specific metrics used for plan evaluation as there are advantages and disadvantages to each depending on the type, size, and location of the target(19). For this reason, visual inspection of the spatial dose distribution and dose-volume histogram (DVH) is often performed clinically to complement quantitative metrics. DVH analysis and measured dose to OARs will therefore also be considered in this work.

### Optimization Platform

In this study we use ROCKET, a stochastic search optimization algorithm which is operated using specialized hardware to solve the combinatorial optimization problem(15, 16). These hardware systems frame the problem in a specific format called Quadratic Unconstrained Binary Optimization (QUBO)(20). QUBO is a powerful formulation for solving combinatorial optimization problems found across a variety of fields including engineering, finance, and government policy(21–24).

An objective function in QUBO format is constructed which simultaneously considers all available parameters to be configured. Penalty terms and coefficients may be added to the function to constrain or prioritize specific output measures of interest. For the purposes of this study, objective functions are constructed for each phase with penalty terms to encourage dose conformity to the target and minimize dose delivered to adjacent healthy tissue. A stochastic Markov Chain Monte Carlo (MCMC) algorithm is run in parallel in order to minimize this function(25). These techniques have been demonstrated to outperform specialized quantum hardware such as D-Wave devices as well as traditional techniques such as simulated annealing and parallel tempering(16, 26).

### Isocenter Positioning

In the first phase of our proposed method, we employ a novel QUBO-based SP algorithm to generate isocenter position coordinates.

A three-dimensional lattice of candidate shot positions is established over the target volume with a binary variable to place or not place a shot at each coordinate. An objective function is constructed which is minimized by the stochastic algorithm to uniformly distribute shots over the target volume. Shots are forbidden outside the target and penalty terms are incorporated to discourage shot placement close to the target periphery in order to prevent excessive dose to healthy tissue. In this phase, shots are defined as spheres with all collimators set to the same size. Shot spheres with dynamic radius are used to accommodate irregularly shaped targets and those with overlapping radii are penalized to avoid large gradients within the dose distribution (*i*.*e*. hotspots).

Once the optimal configuration of isocenter positions is identified, these shot coordinates are held fixed as we proceed to the sector optimization phase where the shot shapes will be finely tuned. It is important to note that this phase allows multiple shots at a single isocenter location. A limitation of the GK hardware is that a single shot weight applies for all sectors in a given shot. While allowing multiple shots at a single isocenter may increase BOT, it also provides more flexibility in adjusting shot shape to achieve a uniform dose distribution across the target.

### Sector Size Optimization

In the SSO phase of the treatment planning procedure, we optimize the sector collimation parameters and weights for each shot. The 192 gamma emitters in the GK helmet are arranged in eight independent sectors of 24 sources. Each sector is equipped with a set of independently controlled collimators to adjust the aperture size for each source. Changing the aperture size will modulate the flux of gamma energy photons through that sector, ultimately adjusting the shape of the dose distribution for each shot. There are four collimator sizes available for each sector: 4mm, 8mm, 16mm, or blocked (0mm). Shot weight (*i*.*e*. exposure time) is also considered as this parameter impacts the overall size of each shot. Upon completion of the SSO phase, the final dose is calculated and the treatment parameters are imported into the clinical software.

The specific objective function we have constructed in this phase simultaneously considers all possible collimator configurations and exposure times (weight) for each shot at their fixed isocenter locations. The individual terms of the function are tuned such that minimizing it results in the maximization of the clinical criteria of coverage, selectivity, gradient index, and BOT. Special consideration is paid in this phase to dose delivered to OARs.

### Minimizing Dose to Healthy Tissue

Among the criteria for evaluating treatment plans is the dose delivered to OARs in the vicinity of the target which often have strict dose restrictions. However, common radiosurgical metrics of coverage, specificity, and gradient index do not directly consider dose to these vulnerable structures. Thus, the sparing of dose to healthy tissue and nearby OARs is included explicitly in the objective function. For the target, a non-overlapping hollow shell called a rind is constructed that conforms to the shape of the target by spatially dilating the target boundary such that the rind volume matches the target volume. By minimizing dose to this shell, the selectivity and gradient index of the dose distribution can be optimized, implicitly minimizing dose delivered to tissue outside the target boundaries. Figure 1 depicts examples of a tumor target, rind, and multiple OARs in the general case and for a specific patient with acoustic neuroma.

**Fig. 1.**
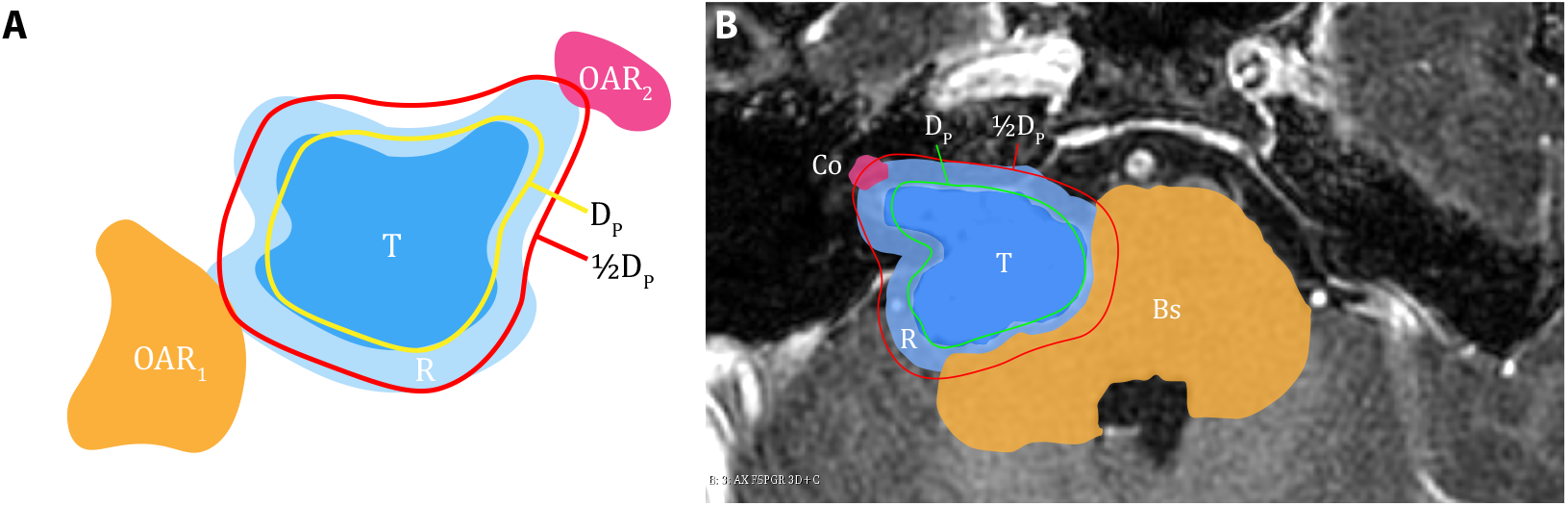
**A)** General diagram of the target T, surrounding rind R, and nearby organs-at-risk OAR_1_ and OAR_2_. Dose delivered to R, OAR_1_, and OAR_2_ is explicitly minimized in the proposed optimization approach. The prescription isodose and 50% isodose lines are indicated by D_P_ and ½D_P_, respectively. **B)** Specific object segmentation in a patient with acoustic neuroma overlaid on a T1-weighted image. OARs are the cochlea (Co) and brainstem (Bs).

Further, dose to nearby OARs is explicitly minimized. Penalty terms are included which consider all OARs together and individually. Including separate terms for each OAR ensures that dose is minimized in those regions regardless of size. For example, the cochlea contributes a relatively small number of voxels to the total volume of OAR in treatments for acoustic neuroma; however, it is highly sensitive to radiation dose resulting in substantial hearing loss or tinnitus(27, 28). Explicit consideration of small OARs promotes safe dose levels to these regions and limits negative treatment side effects.

### Statistical Analysis

A repeated-measures analysis of variance (ANOVA) was carried out to compare treatment metrics across the three planning methods. *P*-values were considered significant at the 0.05 level (false discovery rate, FDR, corrected). A Greenhouse-Geisser correction was used when the data violated the assumption of sphericity. Coverage was scaled to 95% for all plans in this analysis.

## Results

### Overall Results

Optimized treatment plan generation was completed in an average of 51.2 ± 32.3s for SP-ROCKET plans and 27.2 ± 12.3s for Fwd-ROCKET plans. In terms of overall plan quality, optimized GK treatment plans demonstrate equivalent or improved treatment metrics compared to manual forward planning. For this analysis, coverage was set to 95% for all plans.

Target selectivity was increased for all but four subjects (91.8%) in the optimized plans compared to manual forward planning. Average selectivity was significantly improved in SP-ROCKET (0.840 ± 0.058, p=0.001) and Fwd-ROCKET (0.834 ± 0.080, p=0.001) plans relative to forward planning (0.801 ± 0.098). Mean gradient index did not significantly differ between SP-ROCKET (3.046 ± 0.549, p=0.174) and manual plans (2.813 ± 0.194) but was increased in Fwd-ROCKET (3.644 ± 0.945, p< 0.0005) plans. In 30 cases (61.2%), BOT was decreased in optimized SP-ROCKET plans compared to manual forward planning. Average BOT, however, was not significantly different between SP-ROCKET (49.47 ± 18.87min), Fwd-ROCKET (50.00 ± 23.67min), and manual forward planning (56.14 ± 27.77min; p=0.062).

In all but three subjects (93.9%), maximum dose to the cochlea was decreased in the optimized plans compared to forward planning. On average, maximum dose to the cochlea was decreased in both SP-ROCKET (4.20 ± 3.81Gy, p<0.0005) and Fwd-ROCKET (4.41 ± 3.87Gy, p<0.0005) relative to manual forward planning (5.03 ± 4.40Gy). Average values for maximum dose to the brainstem were not significantly different across Fwd-ROCKET (10.44 ± 3.96Gy), SP-ROCKET (10.61 ± 4.59Gy), and manual forward plans (10.28 ± 4.18Gy; p=0.275). Average treatment metrics are shown in Table 1 and depicted graphically in Figure 2.

**Table 1.**
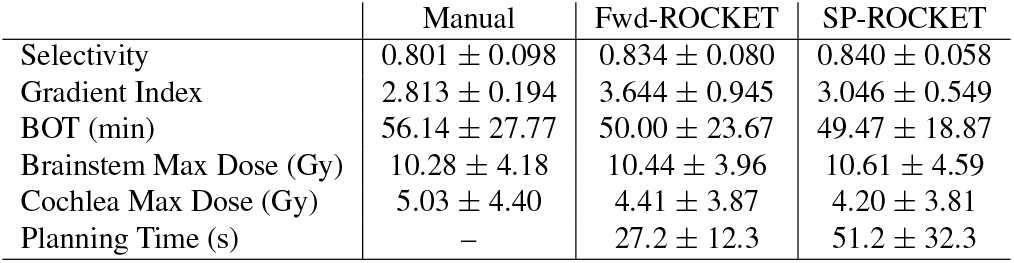
Average metrics for manual forward planning and optimized Fwd-ROCKET and SP-ROCKET plans. Coverage is scaled to 95% for all plans.

**Fig. 2.**
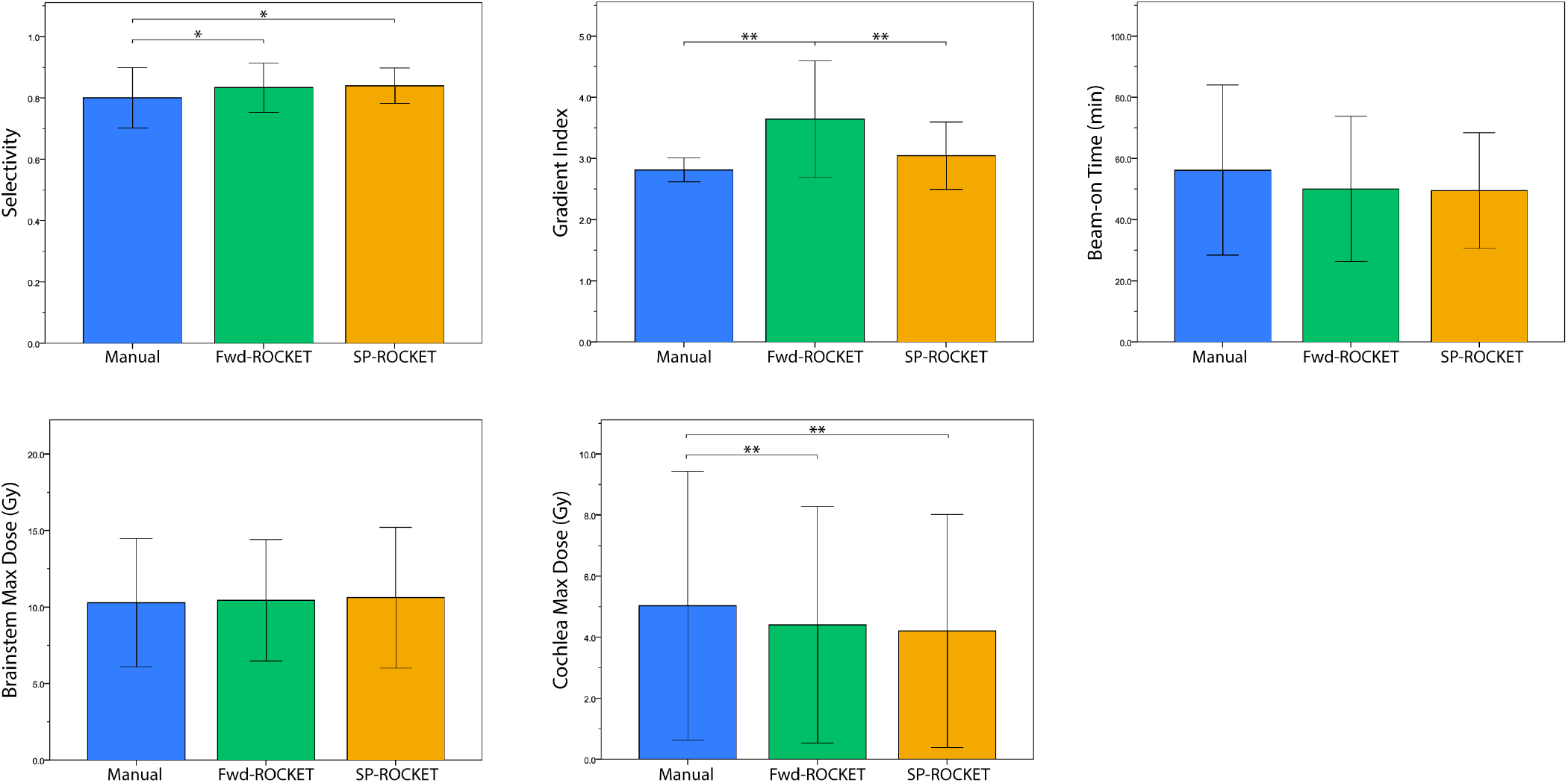
Averaged group data for manual forward planning and optimized Fwd-ROCKET and SP-ROCKET plans. Error bars indicate standard deviation. Significance indicated by *: p<0.001; **: p<0.0005.

### Detailed Case Illustration

We present the results of three subjects in detail which are representative of our proposed planning method. These subjects were all treated for acoustic neuroma and span the range of tumor sizes which might be treated by GK: A) 0.36cm^3^, B) 0.72cm^3^, and C) 5.02cm^3^. Optimized treatment plans for these cases were computed in approximately 70-100s. Table 2 presents treatment metrics for the manual forward plan which was delivered to the patient and the optimized SP-ROCKET plan. These results demonstrate an improvement in treatment quality via increased selectivity and decreased dose to OARs, the brainstem and ipsilateral cochlea.

**Table 2.**
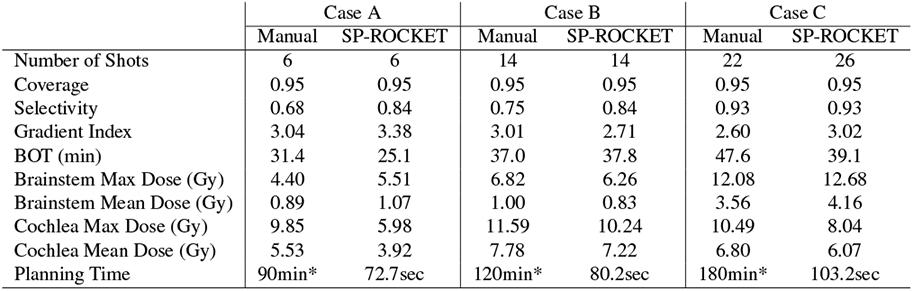
Treatment metrics for sample acoustic neuroma patients with tumor volumes A) 0.36cm^3^, B) 0.72cm^3^, and C) 5.02cm^3^. *: Time estimated by treating neurosurgeon.

DVH plots for these patients are shown in Figure 3, depicting both manual and SP-ROCKET treatment plans. The slope of the SP-ROCKET tumor dose curve closely mirrors the manual reference plan. Decreased rind dose and gradient index in the SP-ROCKET plan indicate a sharper dose drop-off outside of the target. The plots also show substantial decreases in dose throughout the volumes of the brainstem and cochlea, demonstrating enhanced treatment safety due to a decreased likelihood of adverse treatment effects related to healthy tissue exposure.

**Fig. 3.**
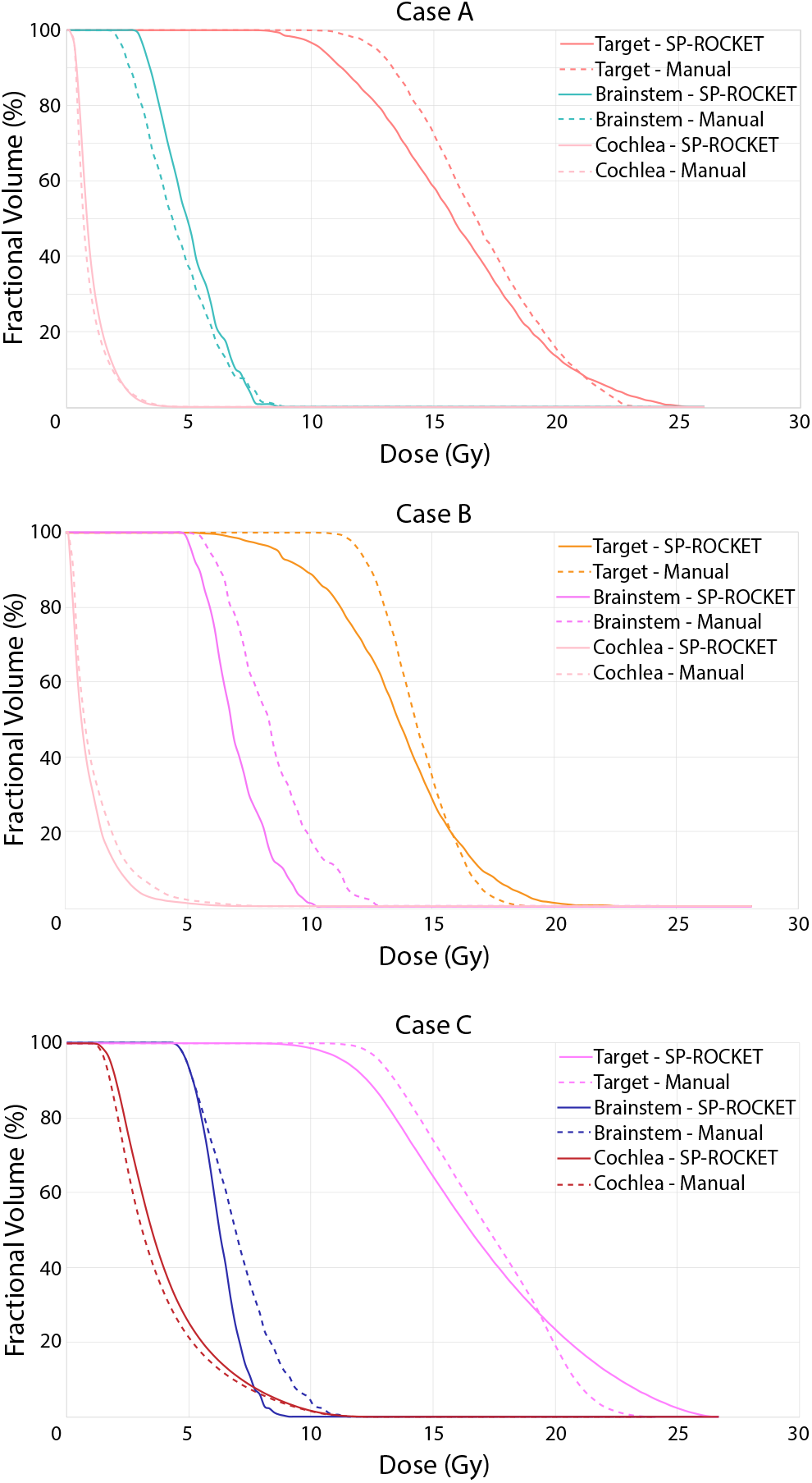
Dose-volume histograms for the acoustic neuroma case illustrations. Manual forward planning is depicted by the dashed line while the optimized SP-ROCKET planning is depicted by the solid line. Dose curves are shown for the tumor, brainstem, and cochlea.

Figure 4 depicts the dose distributions for the acoustic neuroma case illustrations using both manual forward planning and optimized SP-ROCKET planning. The isodose lines demonstrate that there is substantial dose sparing to nearby OARs, particularly the cochlea, while maintaining strong conformity of the prescription dose to the target volume.

**Fig. 4.**
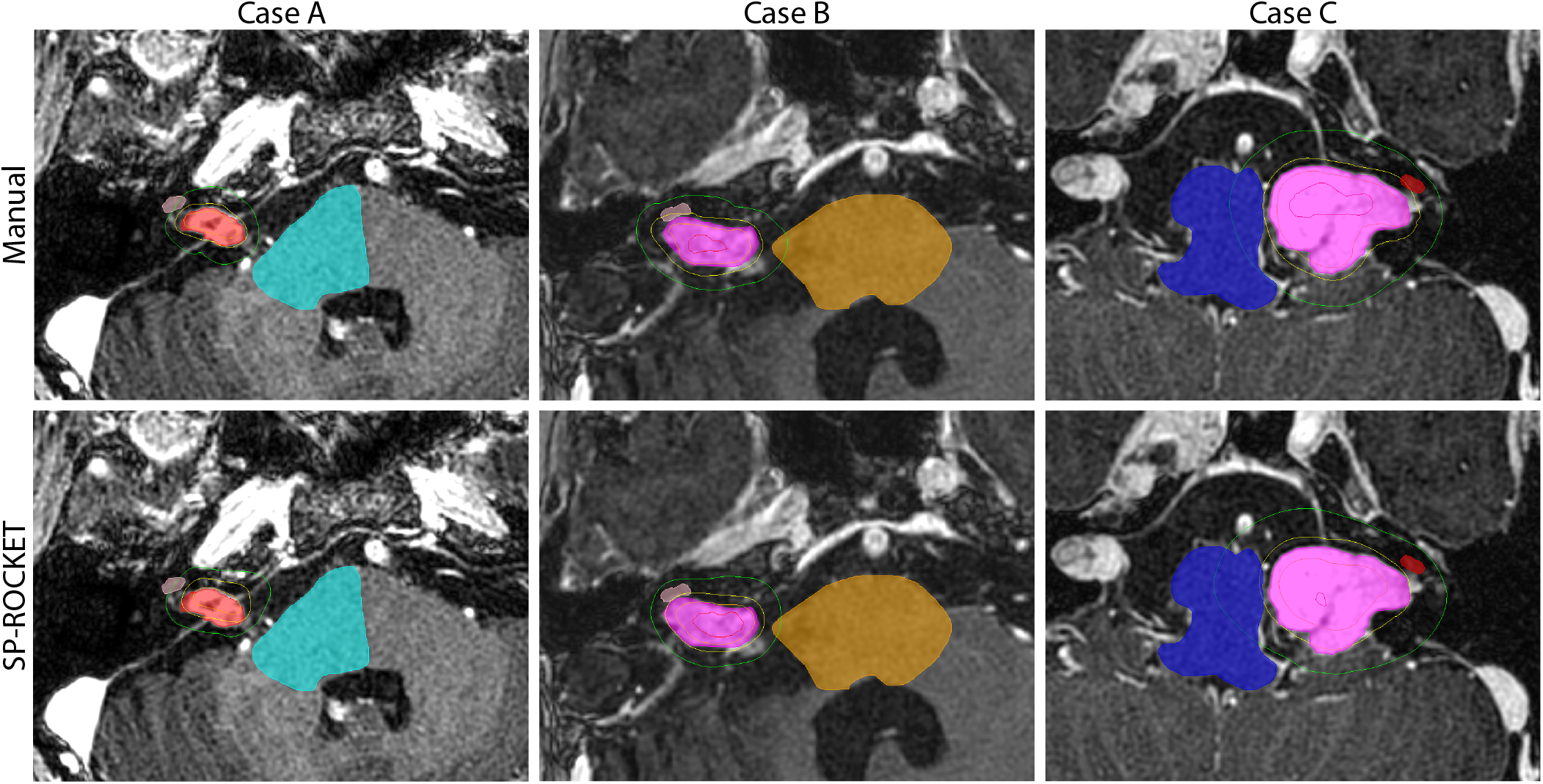
Dose distributions for the acoustic neuroma cases A, B, and C using manual forward planning (top row) and optimized SP-ROCKET planning (bottom row). 5, 10, 15, and 20 Gy isodose lines are shown. Segmentations are shown for the tumor, brainstem, and cochlea. Background is a T1-weighted image in axial view.

## Discussion

In this work we have presented ROCKET, a novel method of automatic GK treatment planning using QUBO-based stochastic optimization. Our results demonstrate that personalized treatment plans can be generated efficiently with quality that is comparable or improved compared to manually generated treatment plans. Specifically, when target coverage is matched between the two approaches, ROCKET treatment plans show improved selectivity and safety through decreased dose to nearby OARs without increasing treatment delivery time. This is accomplished by employing an approach where i) shot isocenter positions are automatically generated and ii) shot shapes are optimized to maximize clinical quality. Automatic GK treatment planning with ROCKET is thus shown to be a robust clinical tool which may enhance the efficiency of clinical radiotherapy treatment planning. While we have demonstrated the use of ROCKET in optimizing GK planning, which tend to be complex and time consuming, this tool is applicable to other types of radiation planning and is non-proprietary to the manufacturers of the GK.

The results presented in this paper are unique as we have demonstrated the ability to generate radiation treatment plans with a substantial time-saving profile while preserving and improving treatment quality. An important feature of our QUBO-based approach is the ability to generate candidate shot isocenters for the optimization. This provides a practical advantage as entire treatment plans may be generated with a single computational platform. A recent work has similarly reported simultaneous optimization of sector duration and isocenters in radiotherapy planning based on an integer programming model(29). QUBO-based solvers, as we use this study, have reported computational benefits relative to these techniques, however(15, 16). Existing approaches to radiotherapy optimization have relied upon non-QUBO SP methods for isocenter determination, including adaptations of the grassfire and skeletonization algorithms(30–33). Others have chosen to focus on shot shape optimization while avoiding manipulation of shot positions(34, 35). The Fwd-ROCKET plans presented in this work demonstrate that our SSO on its own has a positive impact on treatment quality with short computation times. ROCKET may thus be incorporated on its own with other geometry-based, non-QUBO isocenter generation approaches to enhance treatment planning.

The computational techniques used in this study have previously been demonstrated to be effective in other radiotherapy modalities, namely intensity modulated radiation therapy(18), and this work expands on their utility. ROCKET is thus applicable to radiotherapy planning optimization in general and is not restricted a single modality or hardware platform. Reports have also shown these computational approaches to be powerful in other applications such as medical image segmentation(36), finance(37), telecommunications(38, 39), and computer science(40). Other techniques for automatic treatment planning have been reported based on simulated annealing, linear programming, mixed-integer programming, or piecewise penalty methods(29, 34, 35, 41, 42). These models may achieve clinically acceptable plans in terms of quantitative metrics but may have shortcomings in terms of computation time, hardware limitations, or long delivery times. QUBO solvers like ROCKET have previously demonstrated advantages relative to these techniques for solving combinatorial optimization problems such as radiotherapy treatment plan generation(15, 16).

### Limitations

A limitation of this study that may be identified is a disparity in the dose kernels used for calculation of the dose distribution between the manual treatment plans and the DA. The dosimetry software used in the clinical Leksell GammaPlan™ software (TMR10) is proprietary on behalf of the manufacturer and not shared publicly. We did not have direct access to this dose kernel and thus had to reconstruct it locally by generating phantom plans in the clinic with single sectors of each collimator size and summing the contributions of each sector individually for more complex shot arrangements. We believe that our estimation of the dose kernel is robust but recognize the potential for disparity.

Another potential limitation is the choice of metrics used for evaluation of radiosurgery plans. The primary metrics incorporated in our optimization scheme were coverage, selectivity, and gradient index as those are the primary metrics used in the clinic. Other quantitative metrics have been formulated for plan evaluation which may be more appropriate, depending on the diagnosis, size, and location of the tumor(19, 43–45). We note that this limitation applies to both the manual reference plans and those generated by ROCKET.

## Conclusions

We have developed a procedure for generating GK radiosurgery treatment plans automatically using a QUBO-based optimization platform called ROCKET. Personalized treatment plans were generated rapidly and demonstrate equivalent or improved treatment quality compared to manual forward planning. This tool has great potential for the automatic preparation and enhancement of clinical workflow in radiosurgery treatments.

## Data Availability

Data available upon request. Please contact corresponding author.

## ACKNOWLEDGEMENTS

The authors would like to thank Fujitsu Limited and Fujitsu Consulting (Canada) Inc. for providing financial support.

